# When governments spread lies, the fight is against two viruses: A study on the novel coronavirus pandemic in Brazil

**DOI:** 10.1101/2020.10.20.20215962

**Authors:** Priscila Biancovilli, Claudia Jurberg

**Author notes:** Correspondence: Priscila Biancovilli, 7621 Vörösmarty utca. 4, Pécs, Hungary. E.mail. Conflicts of interest: The authors declare no conflict of interest.

## Abstract

**Background:** One of the challenges posed by the novel coronavirus pandemic is the *infodemic risk*, that is, a huge amount of information being published on the topic, along with misinformation and rumours. Around 100 million people in Brazil (50% of the inhabitants) are users of social media networks, and a substantial amount of false information about the disease circulates in these media.

**Objectives:** In this study, we examine the agenda-setting, media frame and content of misinformation published on the topic.

**Methods:** We analysed all pieces of misinformation published by the Brazilian fact-checking service “Agência Lupa”, during six months of 2020. We used content analysis to classify the texts into categories, and three types of rumours were identified: Misleading content; fabricated content; false context.

**Results:** We analysed 232 pieces of misinformation. Most were published on Facebook (76%), followed by Whatsapp, with 10% of total cases. Half of the stories (47%) are classified as “real-life”, that is, the focus is on everyday situations, or circumstances involving people. Regarding the type of misinformation, there is a preponderance of fabricated content, with 53% of total, followed by false context (34%) and misleading content (13%). Wrong information was mostly published in text format (47%). We discuss the influence that misinformation can have on the behaviour of the Brazilian population during the pandemic and how the media’s agenda-setting is influenced by false information published on social media.

**Conclusions:** This study shows that misinformation about COVID-19 in Brazil seem to help establish an agenda-setting in the country, and the media frame is aligned with President Bolsonaro’s political position.

## Introduction

The novel coronavirus (SARS-CoV-2) pandemic generated an avalanche of information that circulates daily around the world. Throughout the first half of 2020, millions of people are or have been quarantined due to the pandemic and suffered negative psychological effects, including post-traumatic stress symptoms, confusion, and anger. Stressors included longer quarantine duration, infection fears, frustration, boredom, inadequate supplies, inadequate information, financial loss, and stigma (Brooks et al., 2020). The Internet is the easiest and fastest source to obtain health information in this context (Starbird et al., 2020). A public health crisis that involves numerous uncertainties requires precise information and answers for the adoption of appropriate behaviours and intelligent decision making (Jang & Baek, 2019).

One of the challenges posed by this new coronavirus is the *infodemic risk*, that is, a tsunami of information about the topic that can also bring rumours and misinformation; with social media, this phenomenon is amplified, and it goes faster and further (Zarocostas, 2020). For this reason, it is important that the population is not only informed in real time but that information needs to be correct and updated, so as many people as possible can act properly to avoid spreading the disease.

### The context of the study: the novel coronavirus pandemic in Brazil

The novel coronavirus in Brazil arrived in a scenario of a conservative, far-right government led by President Jair Bolsonaro, which systematically denies the severity of the SARS-CoV-2 pandemic (Ajzenman et al., 2020). Because of this, the country’s image in facing the pandemic was pointed out as deplorable (Craig, 2020). The attitude of the president and his ministers drew severe criticism from the international press (BBC News, 2020; Financial Times, 2020; The Economist, 2020). Moreover, the World Health Organization (WHO) (Lovelace Jr, 2020) also criticised Brazil’s stance in controlling the pandemic, and for that reason the president threatened to pull Brazil out of the institution (Paraguaçu & Brito, 2020).

The guidelines on wearing masks, social distancing, proper hand washing and the quarantine announced by the World Health Organization and endorsed by the Ministry of Health were routinely questioned by President Bolsonaro, who dismissed two medical ministers in the middle of the epidemic. After appointing a military officer to the post of Interim Minister of Health in May 2020 (Pinto, 2020) the Brazilian government caused, for some days, a blackout in the official data on morbidity and mortality (Leite et al., 2020, BBC News, 2020). Such omission caused a consortium of several broadcasting companies and volunteers to monitor the data daily (Bellini, 2020) in a country with 211 million inhabitants and 5,570 municipalities (IBGE, 2020). In addition to hiding pandemic data in the country, Bolsonaro has encouraged people to go out and even make appearances in stores, markets and public demonstrations on the streets (Ajzenman et al., 2020). As of October, 2020, Brazil had registered more than 5 million cases of the novel coronavirus (Worldometer, 2020a), being the country with the third highest number of cases in the world — the first is the United States and the second is India (Worldometer, 2020b).

In Brazil, about 100 million people are users of social networks (Statista, 2019). This number corresponds to almost half of the country’s total population (IBGE, 2020). Online access to health information has been growing exponentially in the last few years. However, most of the information on the Internet is unregulated, and its quality remains questionable (Cuan-Baltazar et al., 2020).

### Definition and types of misinformation

Misinformation on the Internet started to attract the attention of the media and academics during the U.S. elections in 2016; at that time, the expression “fake news” became increasingly popularised (Vos et al., 2019). Despite the extensive use of this term in the media and in scientific articles, it is considered inadequate to capture the complexity of the information disorder phenomenon (Lazer et al., 2018; Wang et al., 2019; Wardle, 2017). Fake news overlaps with other information disorders, such as misinformation (false, mistaken or misleading information) and disinformation (false information that is purposely spread to deceive or confuse people) (Fetzer, 2004; Lazer et al., 2018).

Even considering these differences, it is not always easy to fit a news item into one of the two categories, because we do not always know if the author of the news had the deliberate intention to deceive, or if he/she really believes in what is being written. For this reason, we follow the same classification as Wang (2019), which uses misinformation as an umbrella term that encompasses all types of false health information, unless the intention to deceive is evident.

The combination of *infodemic* brought about by the novel coronavirus disease situation, with the considerable presence of Brazilians on social networks, many of them without the full capacity to discern the quality of what is published, brings up a potential risk to public health in the country (Uchoa, 2020). The deluge of conflicting information, misinformation and manipulated information on social media should be recognised as a global public health threat (Larson, 2018; Uchoa, 2020). The goal of this study is to analyse the misinformation distributed on social networks under the prism of **intermedia agenda-setting theory** (Vargo et al., 2018) **and media frame theory** (Entman, 1993), using Brazil as a case study.

### Intermedia agenda-setting theory and media frame theory

The original agenda-setting theory (McCombs & Shaw, 1972) was born from a fascinating idea that tries to explain how media plays an important part in shaping political reality. This theory resulted in a significant number of international studies on elections in different parts of the world, such as Argentina, United States, Taiwan, Singapore and Spain, just to name a few (Du, 2013). Over the years, the study of the agenda-setting over time went beyond the political spectrum and sought to understand how the media influences the public debate and the population’s bias in relation to various themes. For example, if the media dedicates a lot of space in their coverage to talk about health, then the population will consider this issue to be of greater relevance. More recently, there are studies that focus on the fake news agenda-setting: even if some audience members are aware that fake news is fake, the rise in coverage can generate an agenda-setting effect; there is also an interplay between different types of media outlets (traditional media, independent media, partisan media, social media users/influencers) in setting each other’s news agenda (Vargo et al., 2018).

About 20 years after the first studies of the agenda-setting, the idea of the media frame was developed by Entman (1993). This term refers to the highlighted selection of certain aspects of reality by the media and, with this, establishes the problematic framing caused by the forms of interpretation, inducing collective thoughts (Entman, 1993). Thus, in addition to the agenda-setting, the media also somehow impose the bias of the news.

According to Entman (1993), there are four actors in this process: the communicator, who chooses what to say, consciously or unconsciously, according to his own experiences; the text, which presents an approach that is determined by keywords, stereotypes, sources of information and phrases that reinforce or not certain aspects of facts or judgments; the receiver, whose interpretations and conclusions are mixed with his experiences; and the culture, defined as the set of frames shared by a given social group.

Using the theoretical framework of intermedia agenda-setting and media frame, this study seeks to analyse how the misinformation disseminated in Brazil about the novel coronavirus guides the discussions within society and affects health behaviours, influencing the progression of the pandemic in the country. To that end, we will try to answer the following questions:

**RQ1: What are the prevalent types, frames and contents of misinformation in Brazil in relation to the pandemic in the novel coronavirus, and what are its main means and formats of dissemination on the Internet?**

**RQ2: Is the misinformation curve on social networks about the novel coronavirus in Brazil directly proportional to the increase in the number of cases in the country, reflecting the growing interest of the population on the topic as the virus spreads?**

**RQ3: In an extraordinary context such as the 2020 pandemic, does the misinformation agenda-setting mostly focus on the disease, or can we observe a heterogeneity of issues addressed?**

## Methods

This is a quali-quantitative exploratory study. We analysed all pieces of misinformation published by the Brazilian fact-checking service “Agência Lupa” (Lupa agency, in English) in the first 27 weeks (six months) of 2020 (from January 1, 2020 to July 4, 2020). Lupa agency was created in 2015 and is the first company specialised in fact-checking in Brazil; the checking is carried out by specialised journalists, based on successful processes implemented by fact-checking platforms such as the Argentine Chequeado and the North American Politifact (Equipe Lupa, 2015).

All news related to the novel coronavirus in the period was organised in an Excel table. For a news item to be considered as related to the topic, the fact-checking text should have at least once the terms “coronavirus” or “COVID-19”. The following aspects of each news item were analysed: a) In what social media has it circulated?; b) What is the content classification and type of misinformation?; c) What is the type of media frame of the false information?; d) Are there recurrent themes in the sample studied?

### Misinformation, content and sentiment analysis of news stories

The nomenclature developed by Wardle (2017) on the different types of misinformation inspired this data analysis. We used the following categories: a) **Misleading content** describes stories which are not entirely false but lead the reader to misinterpret the data; b) **Fabricated content** refers to 100% false pieces of information, with nothing that can be assessed as true; c) **False context** encompasses Wardle’s categories of false context, false connection and manipulated content. The news was classified like this when headlines, visuals or captions do not support the content, or when genuine information (texts, photos or videos) are manipulated; d) **Satire or parody** refers to news that are not meant to be taken seriously, as the main motivation is comical. For this analysis, as in Sommariva et al. (2018) we decided not to include the category e) **imposter content**, when genuine sources are impersonated. This is because the analysis of misinformation producers is not within the scope of this study.

The content analysis of the texts was based on the methodology proposed by Laurence Bardin (1977), which is an inductive process. Firstly, two researchers read in depth (and independently) a sample of 20 news stories. Then, based on the readings, each researcher created a list of categories to describe them. Categorisation followed a semantic criterion: the news was separated according to the theme, and they could not be classified in more than one category. At the same time, the sentiment analysis of each text was also made. Sentiment analysis is the task of identifying positive and negative opinions, emotions, and evaluations (Wilson et al., 2005). Two researchers read all texts and identified the predominant sentiment (positive, negative or neutral) through document-level analysis (Shirsat et al., 2017). The task at this level is to determine the overall opinion of the document. Sentiment analysis at document level assumes that each document expresses opinions on a single entity (Behdenna et al., 2016).

Inter-coder reliability for content and sentiment analysis was 85%. The same process and sample were used to classify the types of misinformation. In this case, inter-coder reliability was 80%. Before classifying the full sample, the authors discussed their experiences and achieved consensus regarding inconsistencies. Then, one author coded the remaining messages.

## Results

### Content and media where misinformation was found

A total of 232 pieces of misinformation were analysed, starting from **week 4** of 2020, when the first fact-checked story was published and finishing at week 27 of 2020. Regarding **RQ1**, we can see a prevalence of certain misinformation contents in Brazil, as shown in **Figure 1**. The stories were classified into seven content categories. Some categories are similar to other studies that analysed discourses in social networks (Biancovilli & Jurberg, 2018; Picanço et al., 2018). We can observe that almost half of the stories (47%) are classified as “real-life”, that is, the focus is on everyday situations, or situations involving people.

**Figure 1.**
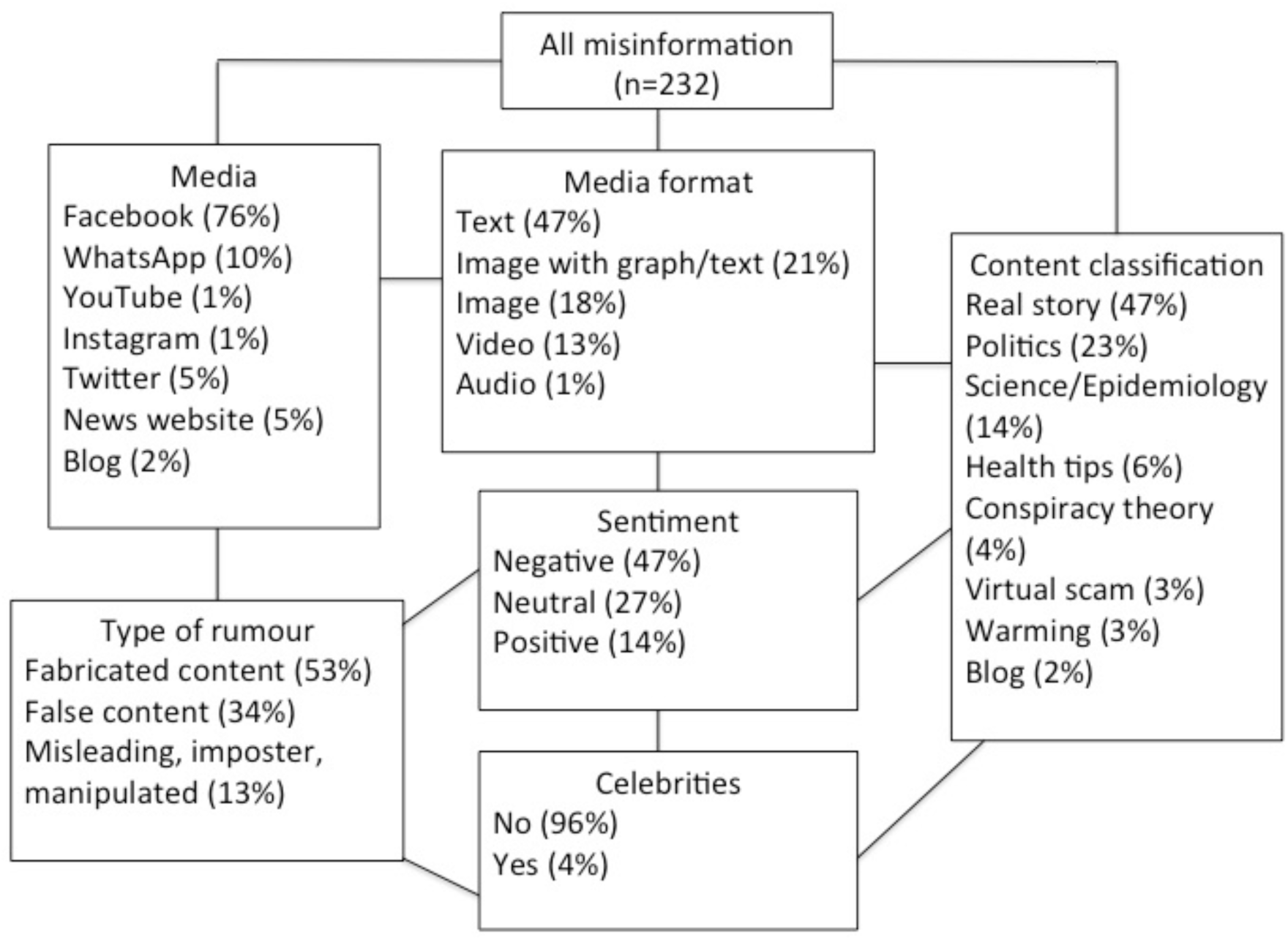
Types and classification of misinformation about COVID-19 in Brazil, between January 1, 2020 and July 4, 2020.

The second most frequent category is “politics”. Here are the stories that focus on politicians, governments, parties, political decisions, aid from governments, laws, decrees, or messages of adulation/adoration aimed at a politician, and they represent 23% of the total amount of registered news. In third place is information on advances in “science and epidemiological data” (14%). Pieces of misinformation related to virtual scams, conspiracy theories and warnings (any kind of warning about what to do or not to do during the pandemic) account for 10% of the total, followed by “health tips” (6%).

Still regarding **RQ1**, most pieces of misinformation were published on Facebook (76%), followed by WhatsApp, with 10% of total cases **(see Figure 1)**. This data sample does not correspond to the ranking of the most used social media platforms in Brazil. According to a recent survey conducted on the habits of Brazilians on social networks (We are Social, 2020), YouTube is the most accessed social media page (96% of Internet users aged 16 to 64 reported using this platform in December 2019); in second place is Facebook (90%), and in third place is WhatsApp (88%), followed by Instagram (79%), Facebook Messenger (66%) and Twitter (48%).

We also observed that the most prominent type of rumour was “fabricated content” (53%), followed by “false content” (34%). It is important to note that all misinformation published on news websites and blogs was also posted on Facebook. When the news appears simultaneously on Facebook and a blog or on Facebook and a news website, it means that there is a link in the Facebook post leading to an external page. In such cases, Facebook serves as a call for the reader to see the full story at the indicated link. In this way, we conclude that 100% of the fact-checked content was published on at least one social network.

Although YouTube is the most popular social media by Internet users in Brazil, only three pieces of misinformation about COVID-19 were found there during this period. Moreover, Instagram and Telegram do not seem to be popular social networks for the spread of misinformation, as only 1% of the total sample was found there.

It is interesting to note that the negative sentiment became predominant throughout the course of the epidemic in Brazil and was mainly associated with “real-life stories” and “politics”. Almost 50% of the information with a negative frame was concentrated on these two themes. Another curious fact was the low use (10/232 or 4%) of celebrities to disseminate false information.

### Misinformation about COVID-19 over time and compared to other topics

Concerning **RQ2**, the first news item recognised by Lupa agency as false in Brazil dates to January 24, 2020 (week 4 of the year). In this week, only one piece of misinformation went through the fact-check process. The peak of fact-checked news occurred in week 14, when 24 pieces of misinformation were published. **Figures 2 and 3** compare the number of diagnosed coronavirus cases per week with the number of pieces of misinformation detected by Lupa agency in the same period. We can see that there is no parallelism between the two phenomena.

**Figure 2.**
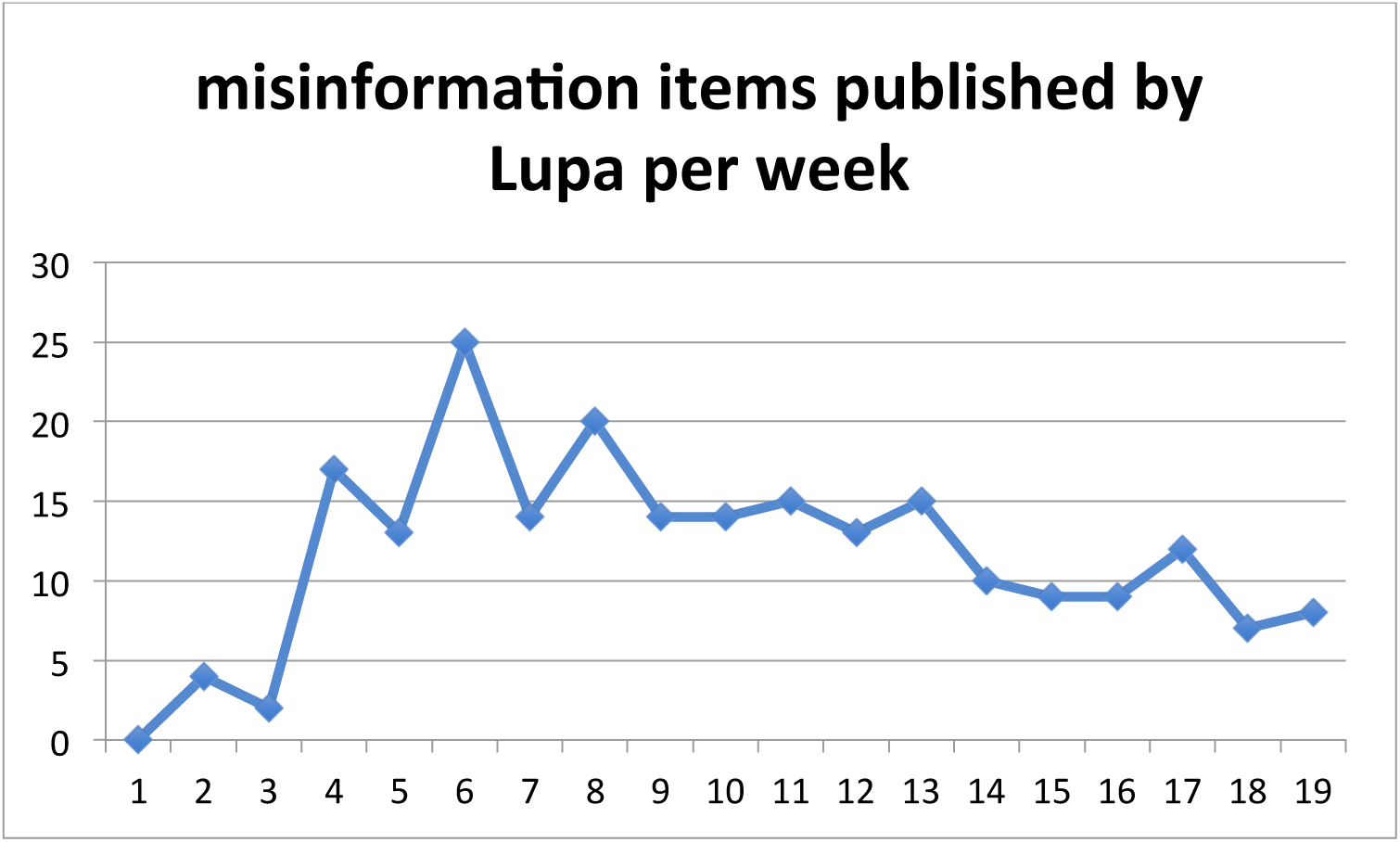
Number of fact-checked misinformation items detected by Lupa agency, since the first case recognised on February 26, 2020 (week 9) until the week 27, in July 4, 2020.

**Figure 3.**
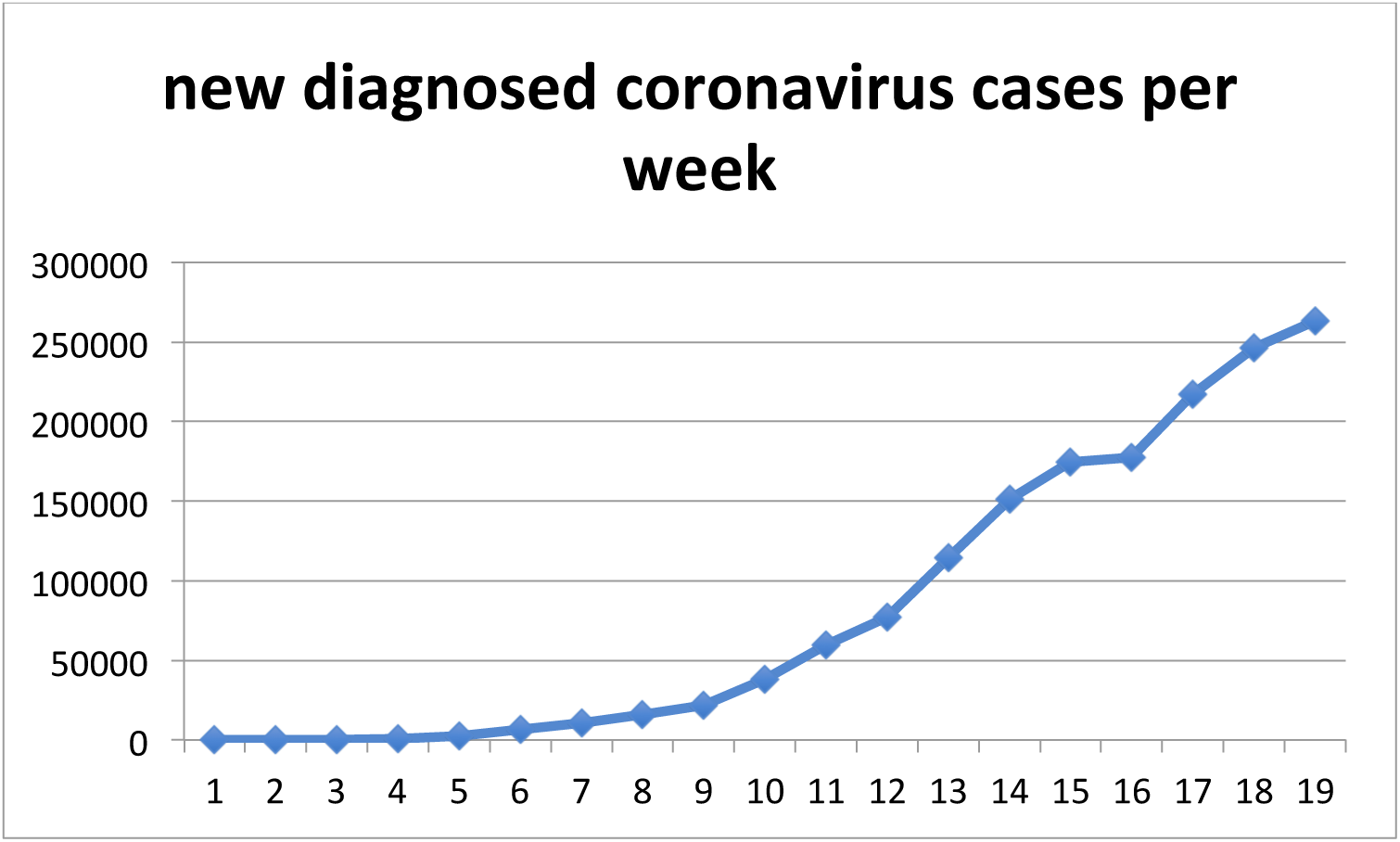
Number of new diagnosed coronavirus cases per week in the same period.

**Figure 4.**
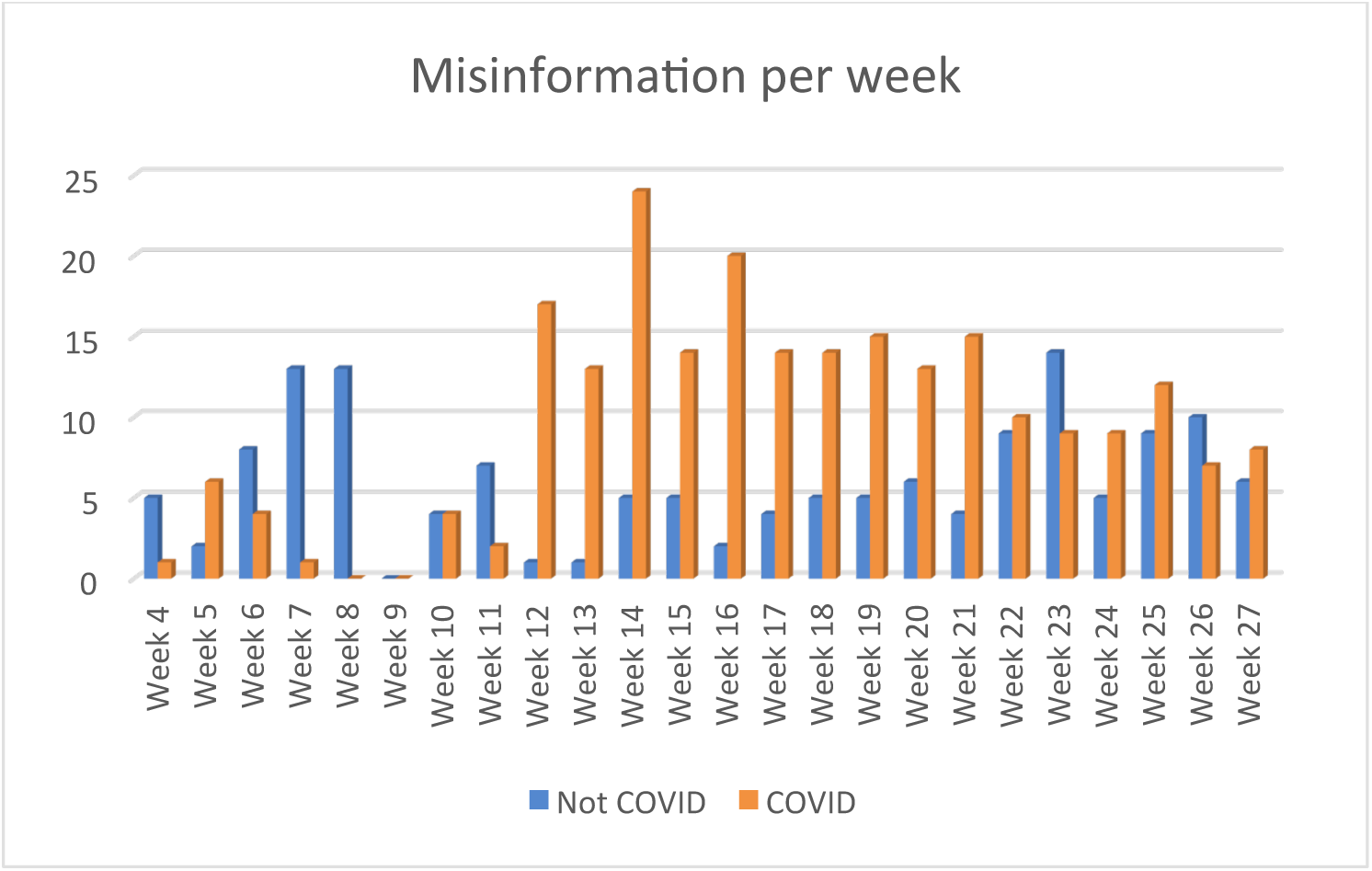
Number of pieces of misinformation that were checked by Lupa agency every week, separated between topics related to COVID-19 and others.

### Agenda-setting of misinformation in a pandemic

Regarding **RQ3**, we noticed that until week 11, the fact-checked content was mostly unrelated to the novel coronavirus. From week 12 onwards, there is a turning point, and in subsequent weeks this issue becomes central to the misinformation agenda-setting. However, in weeks 23 and 26, most of the news items that passed through Lupa agency are not related to the pandemic of the novel coronavirus. Therefore, the misinformation agenda in emergency health situations is not always focused only on the topic. It is important to mention that at the beginning of week 23 (May 31, 2020), there was a series of protests in some cities in Brazil in favour of democracy and against the government of Jair Bolsonaro (Phillips, 2020). For this reason, many false news stories were produced with a focus on demoralising the image of the participants in these protests.

## Discussion

This study explored the spread of misinformation on COVID-19 in Brazil through social media, analysing the stories published by the fact-checking service Lupa agency from January 1, 2020 to July 4, 2020. Although at the end of our sample the pandemic has not yet ended, this study aims to understand the flow of misinformation produced in the first half of 2020 and the agenda-setting, the topics addressed in the country and how the media framing may reflect the positions of social actors and politicians that are relevant in Brazilian society at this time. Brazil is one of the countries that had an exponential increase in the disease case curve during the first half of 2020, and there is not enough literature to try to explain the influence of misinformation on social networks as one of the factors for this problem.

We can see that 100% of the pieces of misinformation in our sample were published on social networks. Even those on blogs and news websites were also posted (via hyperlink) on Facebook, which demonstrates the strength of these new media in spreading information. Social media play a vital role during crises, serving as both a first-hand information channel on the scene as well as a supplementary channel offering specific information demanded by people directly involved in the crisis (Jang & Baek, 2019).

To continue answering RQ3, we can observe that most news detected as misinformation in Brazil (**n=232**) had COVID-19 or novel coronavirus as its main theme, in comparison to news related to other topics (**n=143**). During a pandemic, this type of misinformation can have serious consequences for the population. Mistaken health tips or false scientific and epidemiological data can make people believe that the disease is not so serious, or that a few simple actions (such as drinking tea or gargling) are enough to prevent COVID-19. A study conducted in Northeastern Brazil with 2,259 respondents (Lima et al., 2020) showed that some beliefs of the interviewees are similar to the government speeches denying the seriousness of the pandemic. For the group aged 80 and over, the pandemic in Brazil would be less severe than in many countries; Brazilians would have superior virus protection to people in other countries; and the warm climate of the Northeast of the country would favor the reduction of the pandemic in the region. As this disease is extremely contagious (Kooraki et al., 2020), a less concerned behaviour of the population in relation to it can cause a loosening in measures of social distancing, which contributes to the faster spread of the virus (CDC, 2020) and the overwhelming of hospitals (Powell, 2020).

When we analyse the misinformation agenda-setting in our sample, we observe the attempt to establish some specific themes within society. The themes classified as “real-life stories” and “politics” were the most prevalent in the period. Within these topics, three subjects drew attention: a) Field hospitals supposedly being empty, which proves that the disease is not real (**n=14**); b) People cured by chloroquine and hydroxychloroquine (**n=13**); c) Burial of empty coffins as if they were patients killed by the virus (**n=7**).

The discussion about supposedly empty hospitals gained prominence when the curve of cases of the disease in Brazil began to accelerate, in April 2020 (BBC News, 2020a). Some of the misinformation shows people filming or taking pictures of hospitals with empty receptions. What happens is that many of the hospitals receive only patients referred from other health units, which is why they do not perform emergency care. There are also videos that are contextually false, stating that there are many empty hospital beds, when in fact these videos are old or were filmed in other hospitals in smaller cities. In an official speech after the disclosure of these false news item, Bolsonaro asked supporters to enter hospitals to film whether beds are really occupied (Jucá et al., 2020), which goes against the guidance of doctors due to the risk of contagion. In fact, some of his followers followed his request and invaded hospitals with patients hospitalised for the novel coronavirus, speaking loudly and disrespecting the medical team (Ribeiro, 2020). In addition, in a survey of 833 university students in the health field (Brito Aragao et al., 2020), they stated that the main source of information on COVID-19 was government websites (88.7%), followed by other health institutions (57.3%). That is, despite the government’s denial of the seriousness of the epidemic, official sources remained the most sought after, shaping the agenda setting (McCombs & Shaw, 1972, Vargo et al. 2018) and the media frame (Entman, 1993) of Brazilian news during the pandemic.

Regarding the news involving coffins, one of the pieces of misinformation that most caught the public’s attention was one that said that “coffins of victims of COVID-19 in Belo Horizonte were full of stones”. Another report states that “pits were opened to bury empty coffins in Marabá”. Such misinformation gained prominence on social networks and became a subject in society. For this reason, families began to gather to open sealed coffins with victims of COVID-19, to check if the body they were about to bury was really their family member (Boechat, 2020). In one case, five people were infected due to this action (Pitombo, 2020).

In our sample, 13 of the 232 pieces of misinformation address chloroquine or hydroxychloroquine. All stories treat these drugs as a cure for COVID-19, some with testimonies from celebrities (Tom Hanks’s wife) or anonymous people, and others condemning politicians for not believing in the power of those medicines. Mr. Bolsonaro defended the use of chloroquine against COVID-19 in a pronouncement broadcast on national television in April 2020 (Ricard & Medeiros, 2020). Although there is some evidence about the effectiveness of this substance, data from high-quality clinical trials are still urgently needed (Cortegiani et al., 2020). Another study suggests that high dosage of chloroquine is not recommended for severely ill patients, because of its potential safety hazards (Borba et al., 2020). Due to the sensational and uncontrolled promotion of hydroxychloroquine and chloroquine for the treatment of COVID-19, clinical trials with other more promising drugs and therapies are being pushed aside, because patients refuse to accept them (Ledford, 2020). In other Latin American countries, such as Dominican Republic, clients without a prescription purchase these drugs, as there is a culture of self-medication and lack of governmental regulation on drug use (Tapia, 2020). This same culture exists in all regions of Brazil.

Misinformation propagated in Brazil during the coronavirus pandemic seems to help to establish the agenda-setting in the country, shaping public debates and society’s behaviour (even after fact-checking). The simple fact that journalists dedicate part of their time to address and correct false information, not only at Lupa agency but also in other media, already makes the subject more widespread and, therefore, more debated. A network analysis has found that the sites targeted with the most inbound hyperlinks from fake news networks were mainstream media, social networking sites, and Wikipedia. Few of the targeted sites linked back to the fake news sites (Vargo et al., 2018).

The predominant media frame in misinformation is negationist and endorsed by President Bolsonaro’s speeches, encouraging disrespectful, dangerous and even bizarre behaviour by part of the population. The very fact that the president ignores health recommendations for the pandemic sets a dangerous precedent, causing part of the population to do the same and thus increasing the contagion curve and the number of preventable deaths (Reeves, 2020). Although in the beginning of the pandemic, in March, Brazilians largely adhered to the rules of social isolation and quarantine, over time many people ignored the danger posed by the disease and returned to live their lives normally — even with a growing number of cases and deaths throughout the country (BBC News, 2020c). In June 2020, commerce in the main Brazilian cities reopened; shopping, restaurants and stores started to function normally, and crowds of thousands of unconcerned people in bars were reported by the media (Pedroso et al., 2020; Weir, 2020)

The actions of a political leader are essential to coordinate, organise and ensure that the rules are obeyed by citizens. In a pandemic, the need for a leader who recognises the seriousness of the problem and takes quick action is even greater. A study conducted during the pandemic (Ajzenman et al., 2020) showed that the social distancing measures taken by citizens in pro-government localities weakened compared to places where political support of the president is less strong; they also found evidence that this is stronger in municipalities with a larger proportion of Evangelical parishioners, a key group in terms of support for the president.

Another study that analysed the information disseminated by public health officials during the MERS (Middle East respiratory syndrome) outbreak in South Korea found that less credible information from those professionals led to more frequent use of online news and social media for acquiring information related to the disease. Even though our study did not collect data on the frequency of use of social networks during the pandemic in Brazil, we can observe that there is a relationship between the agenda-setting and the media frame of Brazilian misinformation and the low adherence of the population to preventive care — all this supported by President Bolsonaro’s irresponsible actions. That is, if the country’s leader is not concerned with the severity of the disease, firing two ministers of health in the middle of the pandemic and insisting on chloroquine as a miracle solution, part of the population that supports him feels more confident to return to normality (BBC News, 2020b; Financial Times, 2020; The Economist, 2020). To corroborate this idea, a Brazilian study (Garcia, 2020; Roubaud et al., *in press*) shows a correlation between the preference for President Jair Bolsonaro and the expansion of COVID-19 in certain regions of the country. According to the survey, for every 10 percentage points more votes for Bolsonaro, there is an increase of 11% in the number of cases and 12% in the number of deaths. The researchers characterize the situation as the “Bolsonaro effect” in the spread of the new coronavirus pandemic in Brazil. Other Brazilian studies also reinforce this correlation, showing that whenever the president minimized the pandemic, the rate of social isolation decreases and more people became infected and died (Garcia, 2020).

The guidelines of WHO leaders emphasise the importance of a broad and coherent response from Brazil, especially from governments (at the federal, state or municipal level), to control the pandemic (Ponce, 2020). The lack of a unified response makes the population confused, facilitating denialist attitudes, and neglecting individual actions to protect against the disease.

It is not a coincidence that the misinformation found in this study is similar to the presidential speech. In 2019, an investigation called the “Parliamentary Commission of Inquiry of Fake News” started in the Brazilian Congress, created to investigate the suspected use of false news and misinformation during Jair Bolsonaro’s presidential campaign in 2018. Comprised of 15 senators and 15 deputies, this investigation began with a 180-day period to investigate the creation of false profiles and cyber-attacks on various social networks, with possible influence on the electoral process and public debate (Harris, 2020; Phillips, 2020). In April 2020, the commission was extended and gained another 180 days, which are used to investigate who are the creators and disseminators of disinformation related to the novel coronavirus. There is a strong suspicion that Bolsonaro and his allies (known as the “office of hate”) are participating in schemes for creating false profiles and bots to propagate misinformation against political opponents and contradicting the health measures recommended by the World Health Organization (McCoy, 2020). In July 2020, Facebook has complied with an order by Brazil’s Supreme Court to block the accounts of a dozen top allies of far-right President Jair Bolsonaro (BBC News, 2020d). In other words, the dis- or misinformation agenda-setting and media frame on COVID-19 would not only be endorsed by the president and his allies but also created by them.

## Conclusions and limitations

This study shows that misinformation about COVID-19 in Brazil seem to help establish an agenda-setting in the country, and the media frame is aligned with President Bolsonaro’s political position.

In face of all the challenges discussed in this article, the Brazilian media and science communicators must understand the main characteristics of misinformation in social media about COVID-19, so that they can develop evidence-based content that helps to increase health literacy and modulate the perception of risk. Health educators must have a massive presence in the most popular social media sites with attractive and up-to-date content, as an effort to counteract the spread of misinformation.

As a limitation of our study, we were unable to measure the public engagement of fact-checked stories, to get a more accurate idea of how many people were directly affected by the misinformation. In addition, we have no way of controlling how many people had access to or shared the pieces of misinformation circulating on WhatsApp, due to the private nature of their groups and the lack of data on engagement in this social network. Another limitation is the fact that we used the news verified by Lupa agency only as the basis for this research. As the amount of news circulating about the pandemic is enormous, we have no way of knowing whether the news verified in this period represents the entire sample of misinformation shared in Brazil. Despite these limitations, we believe that this work can offer help so that scientists, journalists and health educators understand the characteristics of the misinformation health agenda-setting during the pandemic and are better able to counter this problem.

## Data Availability

We declare that all data referred to the manuscript entitled When governments spread lies, the fight is against two viruses: A study on the novel coronavirus pandemic in Brazil are avaiable.

## Acknowledgments

This study was supported by the (Brazilian) National Council for Scientific and Technological Development (Conselho Nacional de Desenvolvimento Científico e Tecnológico), and Carlos Chagas Filho Foundation (Fundação Carlos Chagas Filho de Amparo à Pesquisa do Estado do Rio de Janeiro). We would also like to acknowledge Lilian Thomer for developing statistical and mathematical analysis and Mariza Tavares for her insights on misinformation in Brazil.

## Notes

### Competing Interest Statement

The authors have declared no competing interest.

### Funding Statement

This study was supported by the (Brazilian) National Council for Scientific and Technological Development, and Carlos Chagas Filho Foundation. We would also like to acknowledge Lilian Thomer for developing statistical and mathematical analysis and Mariza Tavares for her insights on misinformation in Brazil.

### Author Declarations

The IRB is not necessary in this kind of research.

